# Temperature and relative humidity are not major contributing factor on the occurrence of COVID-19 pandemic: An observational study in 57 countries (2020-05-08)

**DOI:** 10.1101/2020.05.03.20089342

**Authors:** Md. Abdus Sobur, Md. Saiful Islam, Md. Emdadul Haque, AMM Taufiquer Rahman, Md. Taohidul Islam, Antonio Toniolo, Md. Tanvir Rahman

**Affiliations:** Department of Microbiology and Hygiene, Faculty of Veterinary Science, Bangladesh Agricultural University, Mymensingh 2202, Bangladesh; Faculty of Fisheries, Bangladesh Agricultural University, Mymensingh 2202, Bangladesh; Naogaon District Hospital, Naogaon, Bangladesh; Department of Medicine, Faculty of Veterinary Science, Bangladesh Agricultural University, Mymensingh 2202, Bangladesh; Global Virus Network, University of Insubria, 21100 Varese, Italy

**Keywords:** COVID-19, temperature, relative humidity, contact, transmission

## Abstract

The world searching for hope has already experienced a huge loss of lives due to COVID-19 caused by SARS-CoV-2 started in Wuhan, China. There are speculations that climatic conditions can slowdown the transmission of COVID-19. Findings from the early outbreak indicated the possible association of air temperature and relative humidity in COVID-19 occurrence in China. Current study focused on whether climatic conditions (temperature and relative humidity) are having any influence in the occurrence of COVID-19 when the outbreak has been classified as pandemic. To determine the effect of daily average temperature and average relative humidity on log-transformed total daily cases of COVID-19, polynomial regression as a quadratic term and linear regression were done. Linear regression analysis was also carried out to explore the same effect on selected countries. Present study observed no correlation between the climatic conditions (the daily average temperature and relative humidity) and the number of cases of COVID-19. Similar result was found in relation between daily average temperature and average number of cases per day in country-wise analysis. However, about 93.5% cases of COVID-19 occurred between 1°C to 16°C and the average number of cases per day was lower in high temperature country than low temperature country with exceptions. The minimum effect of summer temperature may not be effective to control the pandemic rather need to apply the control measures of COVID-19.

## 1. Introduction

COVID-19 caused by SARS-CoV-2 is an enveloped, positive sense single stranded RNA virus of the family *Coronaviridae*, a major pathogen of respiratory illness (Pal et al., 2020). Since the outbreak of COVID-19in Wuhan, China on the late December, 2019 (Lu et al., 2020), it has rapidly spread throughout China and all over the world (Gorbalenya, 2020; Arab-Mazar et al., 2020; Chen et al., 2020; Huang et al., 2020; Wang et al., 2020a; Holshue et al., 2020; Wang et al., 2020b). The major contributing factor for this new epidemic was the importation of COVID-19 cases from epidemic countries. So far, more than 200 countries and territories have been affected with 1,395,136 confirmed cases and 81,580 deaths, togetherwith cessation of normal social operations and massive economic loss (WHO, 2020a). Earlier, WHO, observing the current situation across the world, declared COVID-19 as “a pandemic” (WHO, 2020b).

The experience of SARS and MERS outbreak differs with COVID-19 in extensiveness of spread, and death rate but finds similarity in sign-symptoms and transmission pattern (Wang et al., 2020c). Seasonality of respiratory viral disease is an old theme that recognized winter as an important factor of influenza like diseases transmission (Sobsey and Meschke, 2003; Wolkoff, 2018). The transmission of virus is under the influence of climate conditions e.g., temperature and relative humidity (Hemmes, et al., 1960, Dalziel, et al. 2018). Several studies found droplet, aerosol, and direct and indirect contact can act as the major factors in the transmission of SARS like viruses (Peng et al., 2020; Rodríguez-Morales et al., 2020; To et al., 2020). Survivability of SARS virus in environment and surfaces is associated with temperature and relative humidity(Chan et al., 2011). High temperature reduces aerosol but cannot inhibit contact transmission immediately (Lowen et al., 2008). SARS-CoV-1 and SARS-CoV-2 are able to survive in air and surface for sizable time periods, thus enabling the virus more capable to affect human (Van Doremalen et al., 2020). Survival of SARS-CoV-2 in air and surfacesassociated with climate with is still unclear.

Although some studies found relationships of climatic conditions with the occurrence of COVID-19 cases (Wang et al., 2020d, Wang et al., 2020e, Zhu and Xie, 2020), they had limited option to include high temperature country as the outbreak was manly circulating in temperate zone during the early stage of outbreak. Therefore, this study aimed to understand the possible role of temperature and relative humidityin the occurrence of COVID-19 in 57 countries from tropical and temperate zone.

## 2. Materials and methods

### 2.1. Data

Countries having at least 500 cumulative cases by March 28, 2020 were selected forstudy. Once a country reported at least 10 cases on particular day was considered for data recordingfor that day. Date wise metrological parameter, like temperature and relative humidity were obtained from 57 countries to find out a possible link between COVID-19 cases and metrological data. The highest and the lowest temperature were recorded and the average of these two data was considered as the daily temperature. The daily relative humidity was recorded as AM (Before mid-day) and PM (After mid-day). The average was considered as the daily relative humidity.

### 2.2. Source of Data

We extracted information from national and international reports of the outbreak and metrological data from relevant websites. The daily case reports were documented from “WHO”, “Worldometer”, and “Kaiser Family Foundation”websites (WHO, 2020c; Worldometer, 2020; KFF, 2020). The daily temperature and relative humidity data were recorded from “The Weather Channel”, “Timeanddate” and “Accuweather” websites (The Weather Channel, 2020; Timeanddate, 2020, Accuweather 2020). All the data taken for this study were from January 9, 2020 to March 25, 2020.

### 2.3. Statistical Analysis

Natural Log-transformation was performed on total number of daily COVID-19 confirmed cases of 57 countries. Polynomial regression as a quadratic term was done to find out the relationship between daily average temperature and log transformed daily cases of COVID-19 as we observed a non-linear relationship during initial data exploration (Bradley and Srivastava, 1979). However, the linear regression was used to find out the effect of daily average relative humidity on COVID-19 cases (Matthews, 2014). For a certain country, the natural log of the average number of cases per day was used, and the linear regression was done to find out the effect of temperature and relative humidity on the occurrence of COVID-19 cases.

## 3. Results

The data on average temperature, average relative humidity, and log transformed confirmed cases per day of COVID-19 in 57 countries are listed in Supplementary Table. Here, we included 444,405 cases from 57 countries (January 9, 2020 to March 25, 2020). The daily average lowest and highest temperature was respectively −5°C in Finland and 29.9°C in Thailand. Percent relative humidity varied from 30 in Saudi Arabia to 78.8 in Indonesia. Data suggest relative humidity was fluctuating with large deviation in every country. Except China (January, February, March), Italy (February, March), Iran (February, March), South Korea (February, March), France (February, March) and Japan (February, March) all other countries had COVID-19 cases in March. In China, the early outbreak was in January when average cases per day were lower than in February and March. Similarly, Italy, Iran, South Korea, France and Japan had lower average cases per day in February compared to March. Among the countries, Italy had the lowest and highest average cases per day respectively in February (average monthly temperature 7.5°C) and March (average monthly temperature 9.4°C).

In polynomial regression as a quadratic term, no correlation (R^2^ = 0.031) between the daily average temperature and the number of cases of COVID-19 was found. The analysis showed that lgN of cases increased as the average temperature rose and started to decline moderately when temperature reached the peak (Fig. 1). The analysis showed that 98.8% proportionate cases occurred between −0.5°C to 30°Cand, more restrictively, 93.5% cases occurred between 1°C to 16°C (Fig. 1). The average relative humidity was also not correlated (R^2^ = 0.0272) with the occurrence of COVID-19 in linear regression (Fig. 2). However, most of the low temperature countries had relatively higher average number of cases per day than high temperature countries (Fig. 3). There were also exceptions. For country-wise analysis, average monthly observations were used for each country. In linear regression country-wise analysis revealed no correlation (R^2^ = 0.0602) between temperature and occurrence of COVID-19 cases per day (Fig. 3).

**Figure 1.**
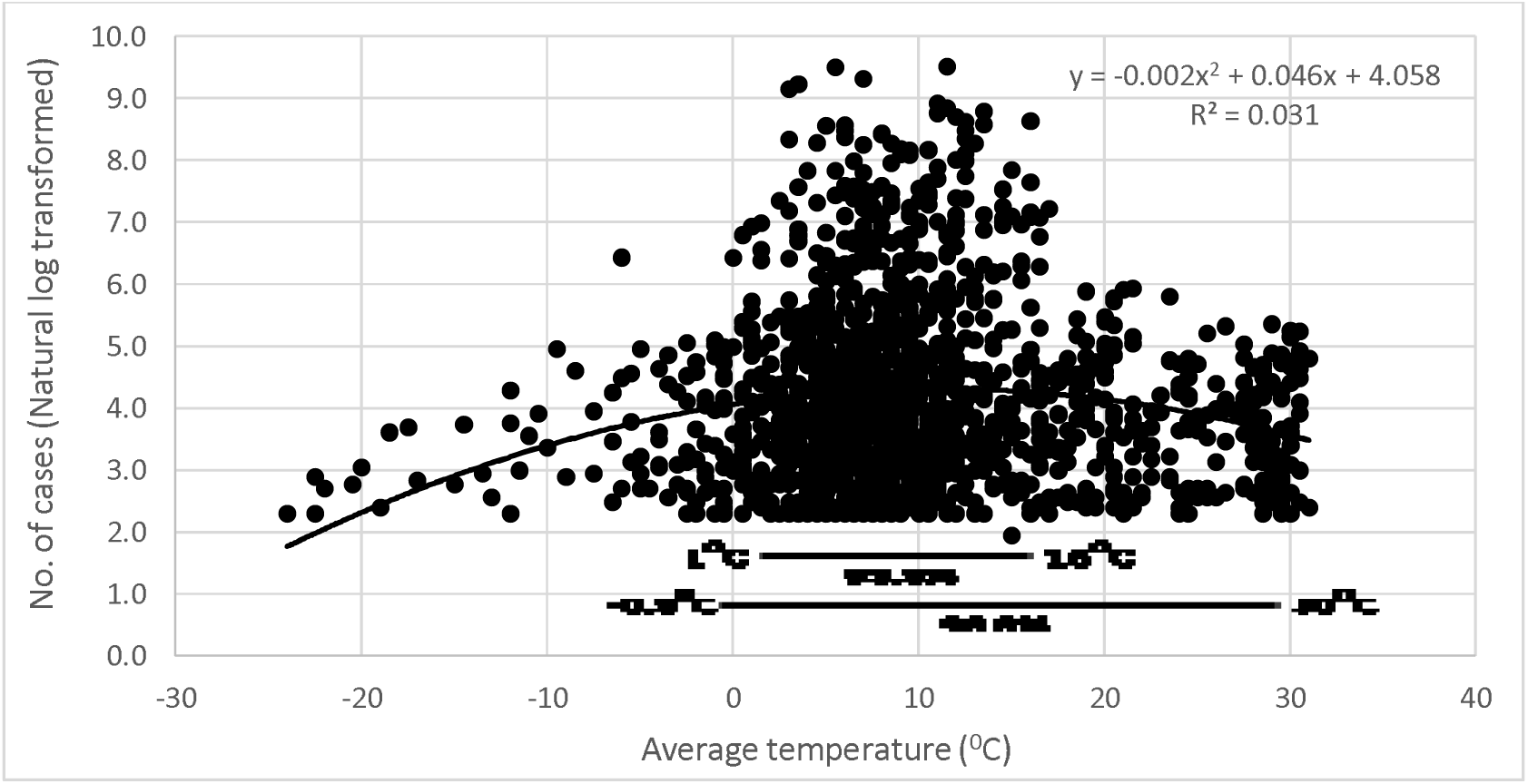
The relationship between average temperature and COVID-19 cases (natural log transformed) in 57 countries

**Figure 2.**
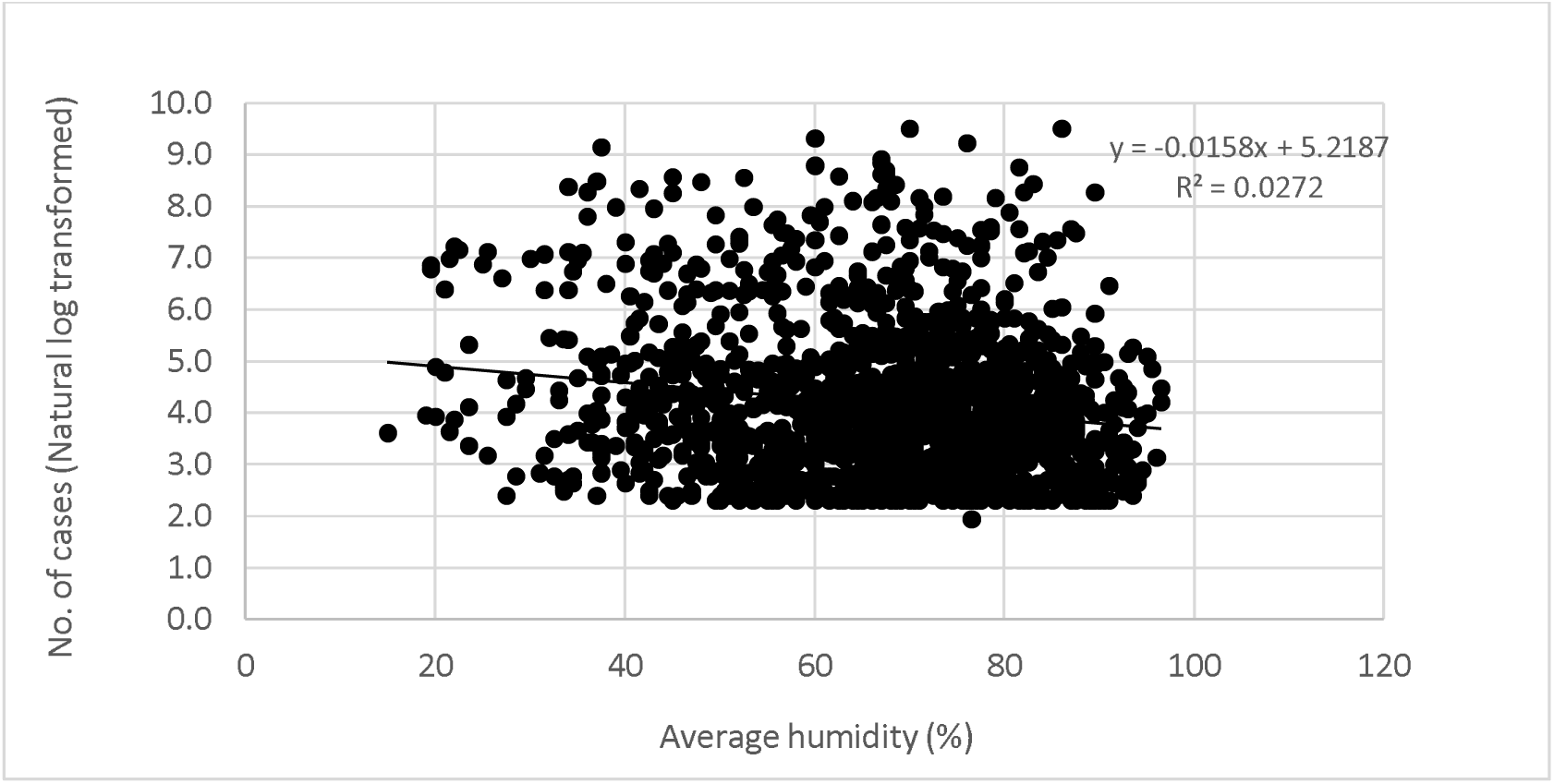
The relationship between average relative humidity and COVID-19 cases (natural log transformed) in 57 countries

**Figure 3.**
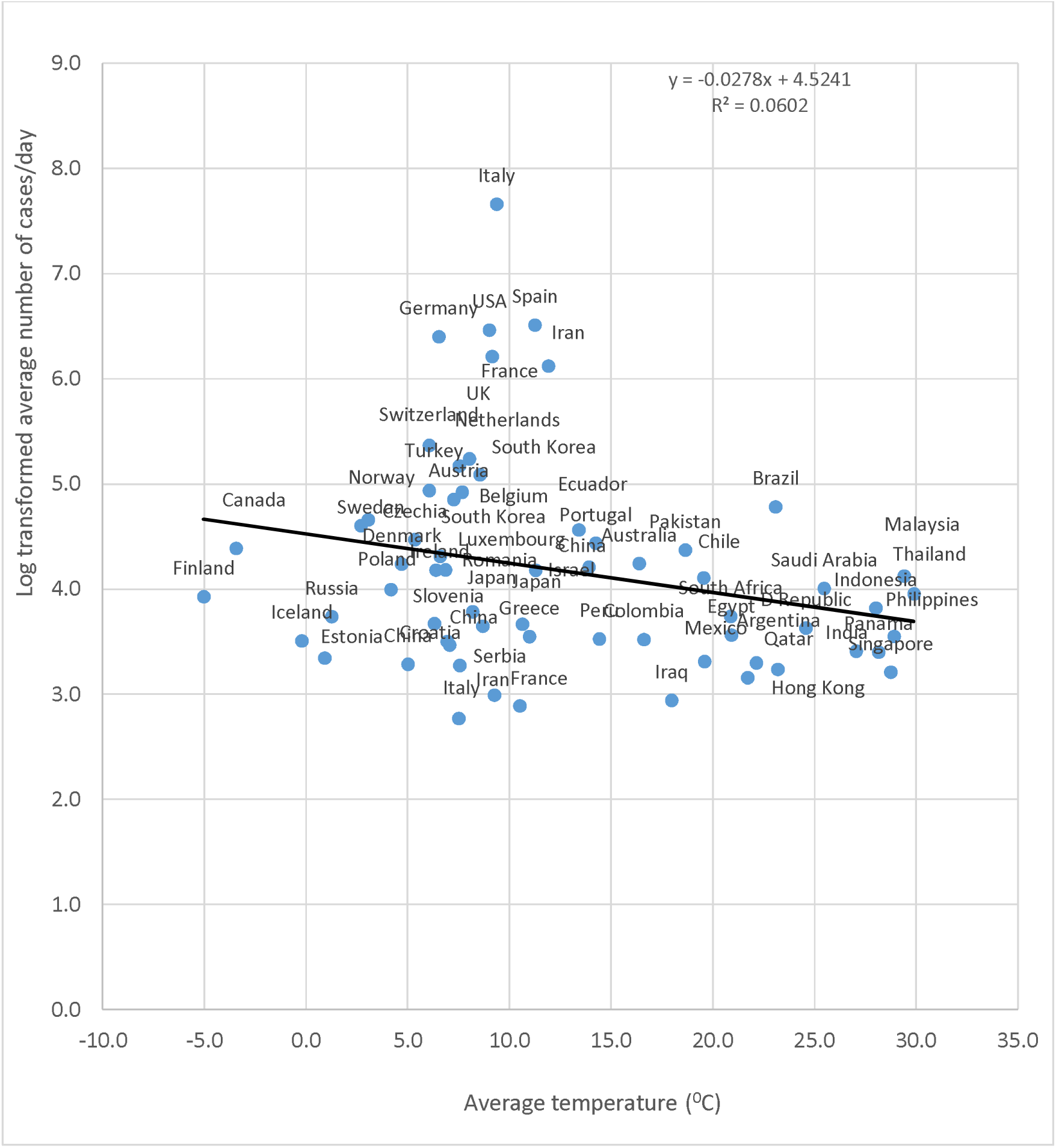
Country-wise log transformed average cases of COVID-19 per day v.s. average temperature in 57 countries

## 4. Discussion

This is the first study that consider the pandemic situation (57 countries including high and low temperature countries)to draw conclusions on possible association of temperature and relative humidity with COVID-19 cases. Therefore, the findings of this study demand importance. No correlation between the daily average temperature and the occurrence of cases of COVID-19 was observed. Earlier studies found correlation between temperature and the occurrence of COVID-19 (Wang et al., 2020d, Wang et al., 2020e, Zhu et al 2020). Wang et al., (2020d) showed that most COVID-19 cases in China occurred between 0°C to 10°C. Another study analyzed data (January 20, to February 4, 2020) of COVID-19 cases and revealed that the majority of cases occurred with temperature around 10°C (Wang et al., 2020e). But all these studies obtained data from the early stage of the outbreak, at best till February when most of the affected countries were in winter. In our study, temperature varied from −24°C to 30.5°C in 57 countries. The huge variation of temperature in analysis found no correlation with COVID-19 cases. We found 93.5% COVID-19 cases were between 1°C to 16°C as most of cases were from China, Italy, USA, Iran, Germany, France and Spain where the duration of outbreak is prolong (Supplementary Table).

In country-wise analysis, no correlation was observed between average daily temperature and average per day cases of COVID-19. However, in general, countries having high temperature experienced relatively lower occurrence of COVID-19 (Fig. 3) with exceptions. This is may be due to high temperature countries are at early stage of outbreak. With the progress of outbreak the number of cases may increase exponentially if proper control measures are not taken. The transmission was exponential in some developed low-temperature countries like USA, Italy, Spain, Germany, and France where large scale community transmission was reported (WHO, 2020a). On the other hand, similar countries like South Korea, Japan took steps during early stage of the outbreak being quite successful in limiting virus transmission (Financial Times, 2020). The average daily temperature in Hong Kong (21.7°C) and Brazil (23.1°C) was quite similar, but Brazil had relatively higher number of daily average cases (Supplementary Table). These findings suggest all courtiers for applying proper control measures. However, these exceptions can be interpreted taking into consideration that virus transmission in an outbreak largely depends on the interventions taken by each particular government. In addition, duration of outbreak, population density, other metrological variables, level of awareness in people, and medical facilities are not same for all countries. Among so many factors, transmission of COVID-19 virus from asymptomatic is carrier very important. A study has been carried out on the passengers of Diamond Princess cruise ship, where 50.47% COVID-19 patients were asymptomatic carrier (Mizumoto et al., 2020). Additional study is required to consider many more variables. A variable relative humidity was considered in our study. Though Wang et al., (2020d) found a correlation between relative humidity and the occurrence of COVID-19 cases, we could not confirm this association. A recent study in China Jakarta, Indonesiaalso showed no effect of relative humidity on COVID-19 occurrence (Tosepu et al., 2020). This observed variation in results may be due to a variation in the investigated time periods and data size in the different studies. In addition we observed inconsistent relative humidity in most of the countries in our study.

The stability SARS virus was studied in laboratory, at 22-25°C SARS-CoV is stable up to five days on plastic surface while rapidly losing activity at 38°C or more (Chan et al., 2011). SARS-CoV-2 survive in air (3 hours), on plastic and stainless steel surfaces (up to 72 hours) thus contact transmission is facilitated (Van Doremalen et al., 2020), but investigations of this type associated temperature and humidity are scarce for SARS-CoV-2. Preliminary evidences indicate that high temperature affect virus transmissibility through droplet and aerosol (Lowen et al., 2008), but contact transmission cannot be ignored (Van Doremalen et al, 2020). Therefore, “high temperature” itself is not enough to combat the pandemic. The primary epicenter in Wuhan, Chinaalready has proved that the virus is controllable in a low-temperature region (WHO, 2020d). We suggest that all countries should follow control measures similar to those adopted in China, taking into account that temperature increments will not be enough.

## 5. Conclusions

To the best of our knowledge, this is the first study describing temperature and relative humidity has no major effect to describe on the current COVID-19 pandemic. More studies needed to which variables are contributing most. Current study suggests all countries not to depend on summer rather to implement possible all control measures of COVID-19.

## Data Availability

Not applicable.

## Author’s contribution

MTR and MAS designed the study. MAS, MSI and MEH collected data. MAS, MEH and MTI analyzed and interpreted the data. MAS, MSI, AT, MTI, AMMTR and MTR drafted the manuscript. MAS, MTI, AT, AMMTR and MTR critically reviewed and updated the manuscript to its final version. The final version of the manuscript was approved by all authors.

## Supplementary Table legend

**Supplementary Table 1.**
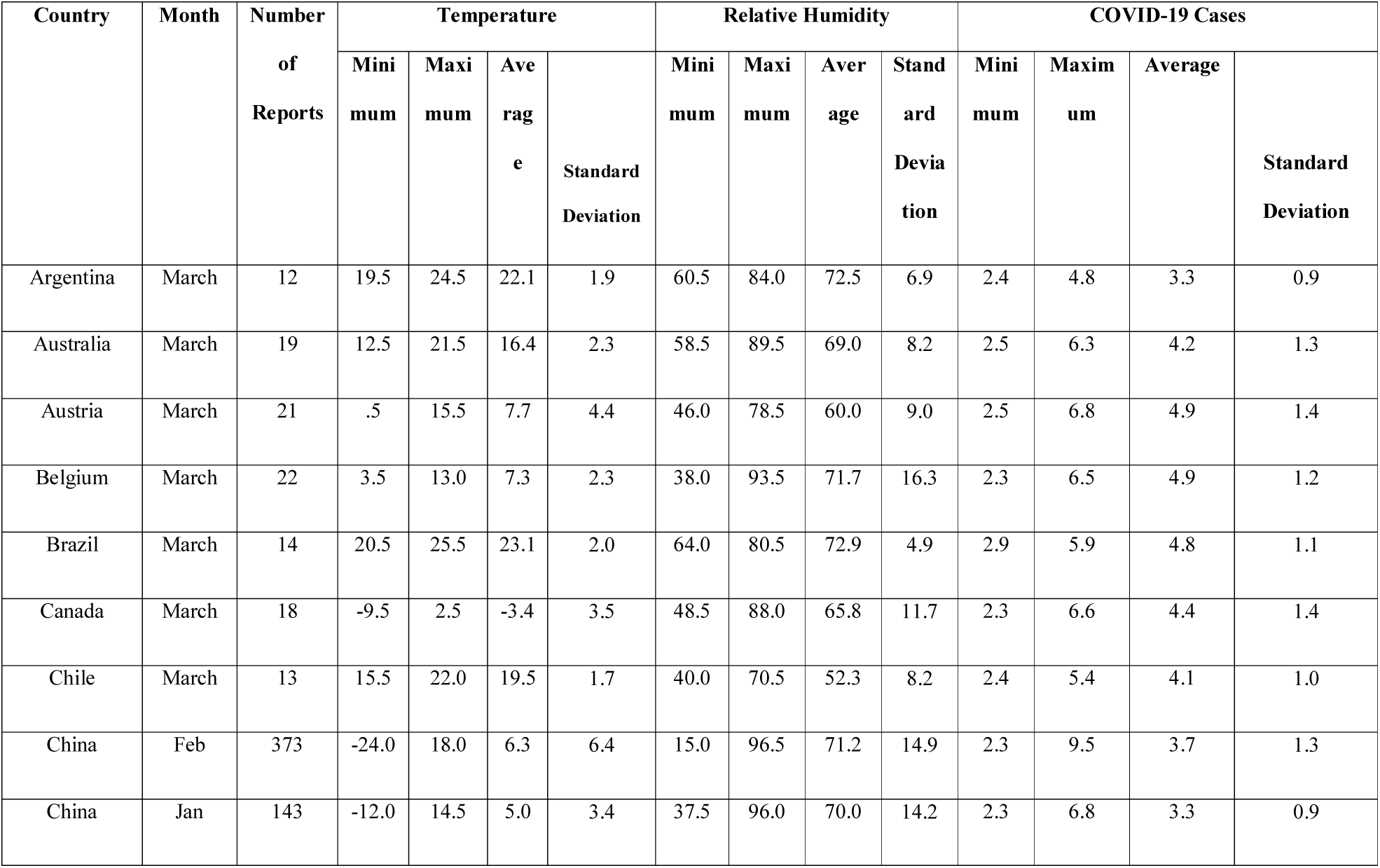
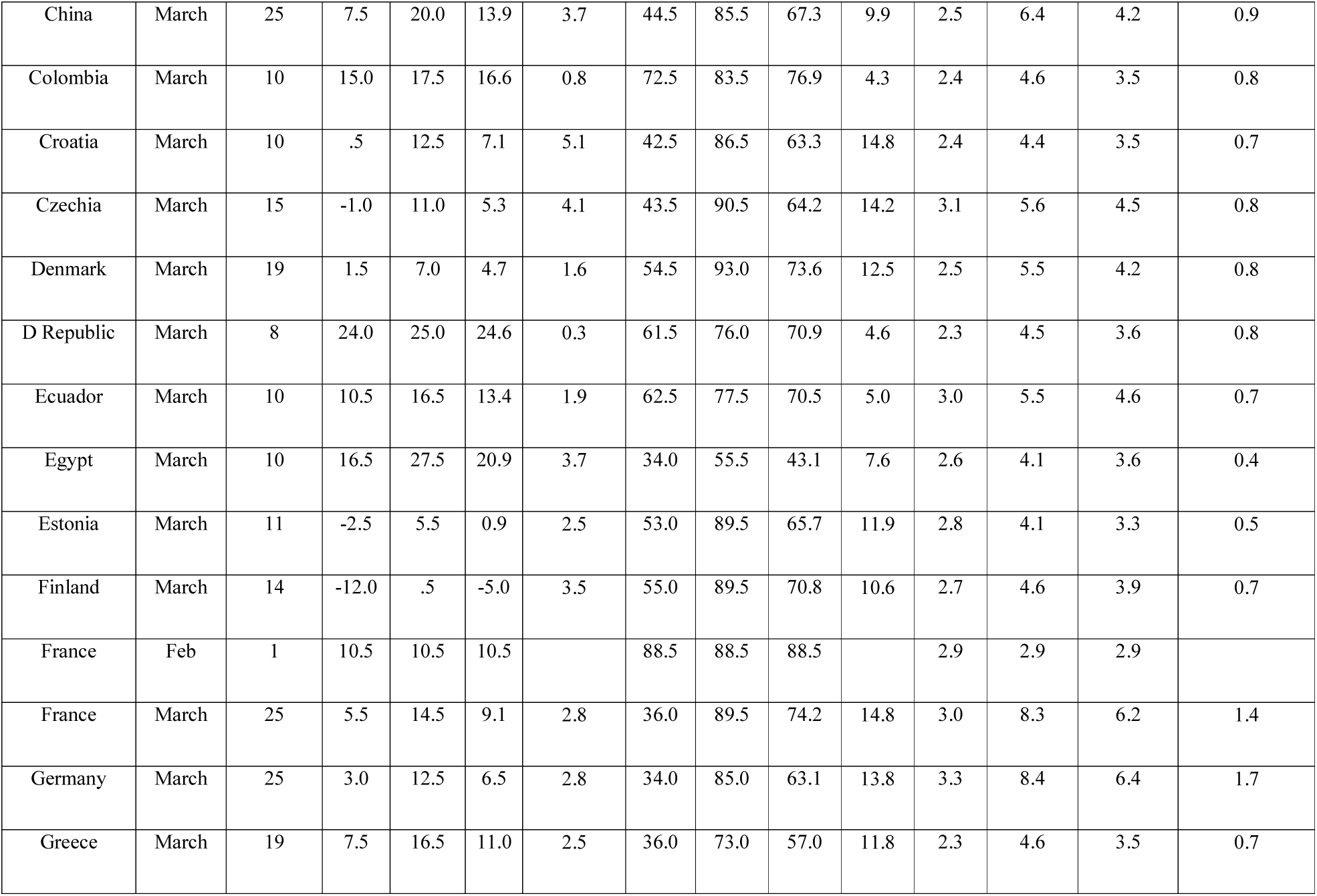
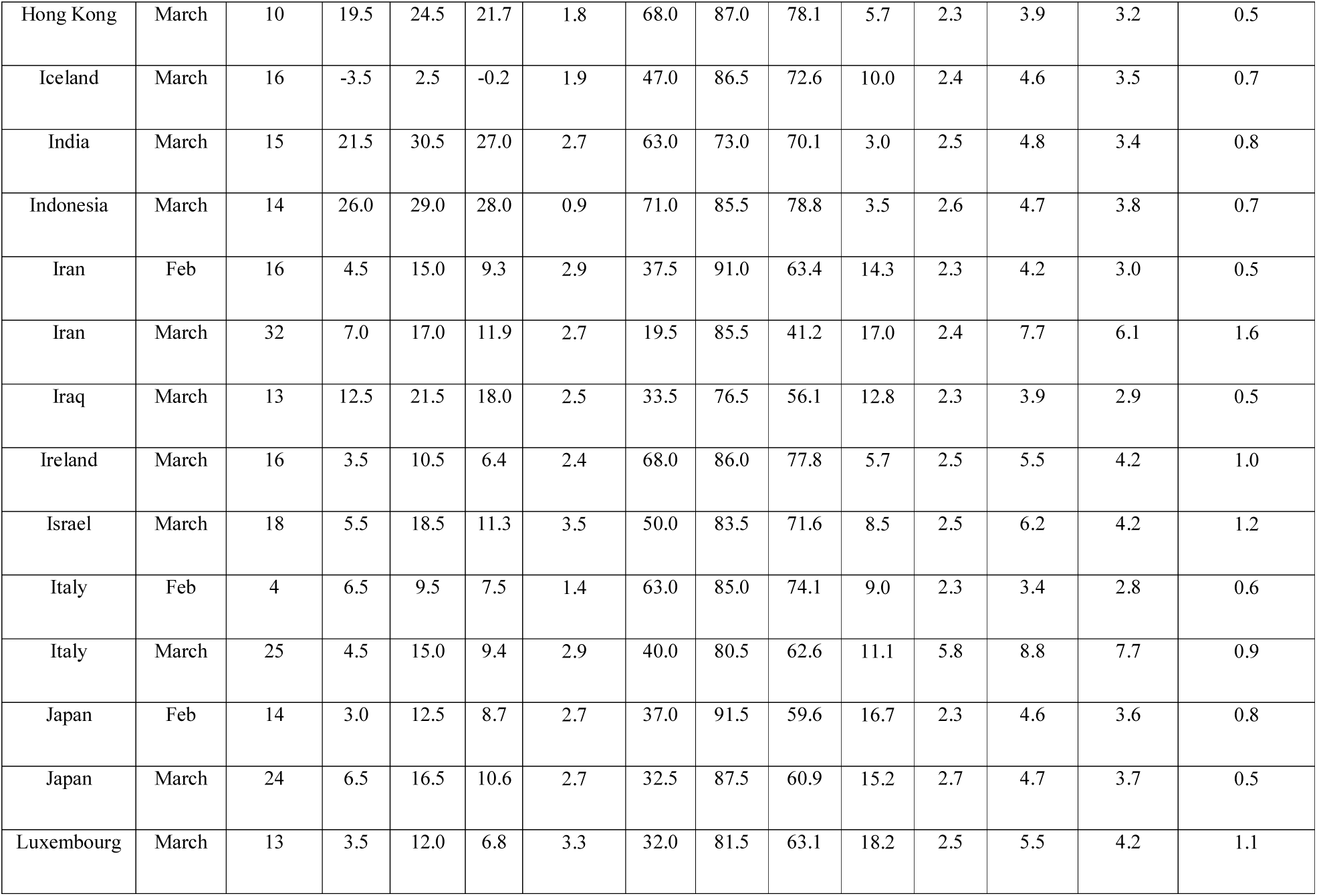
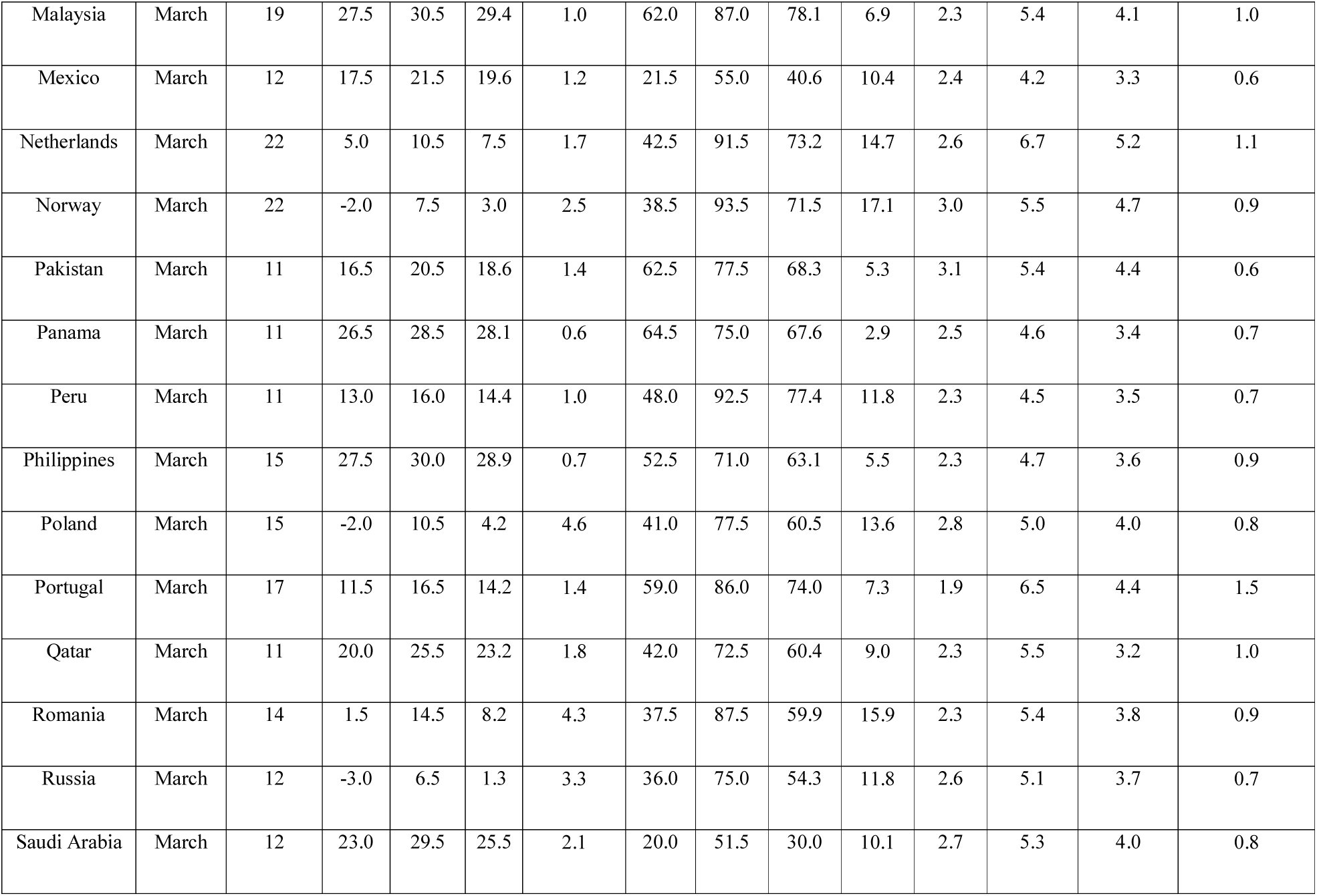
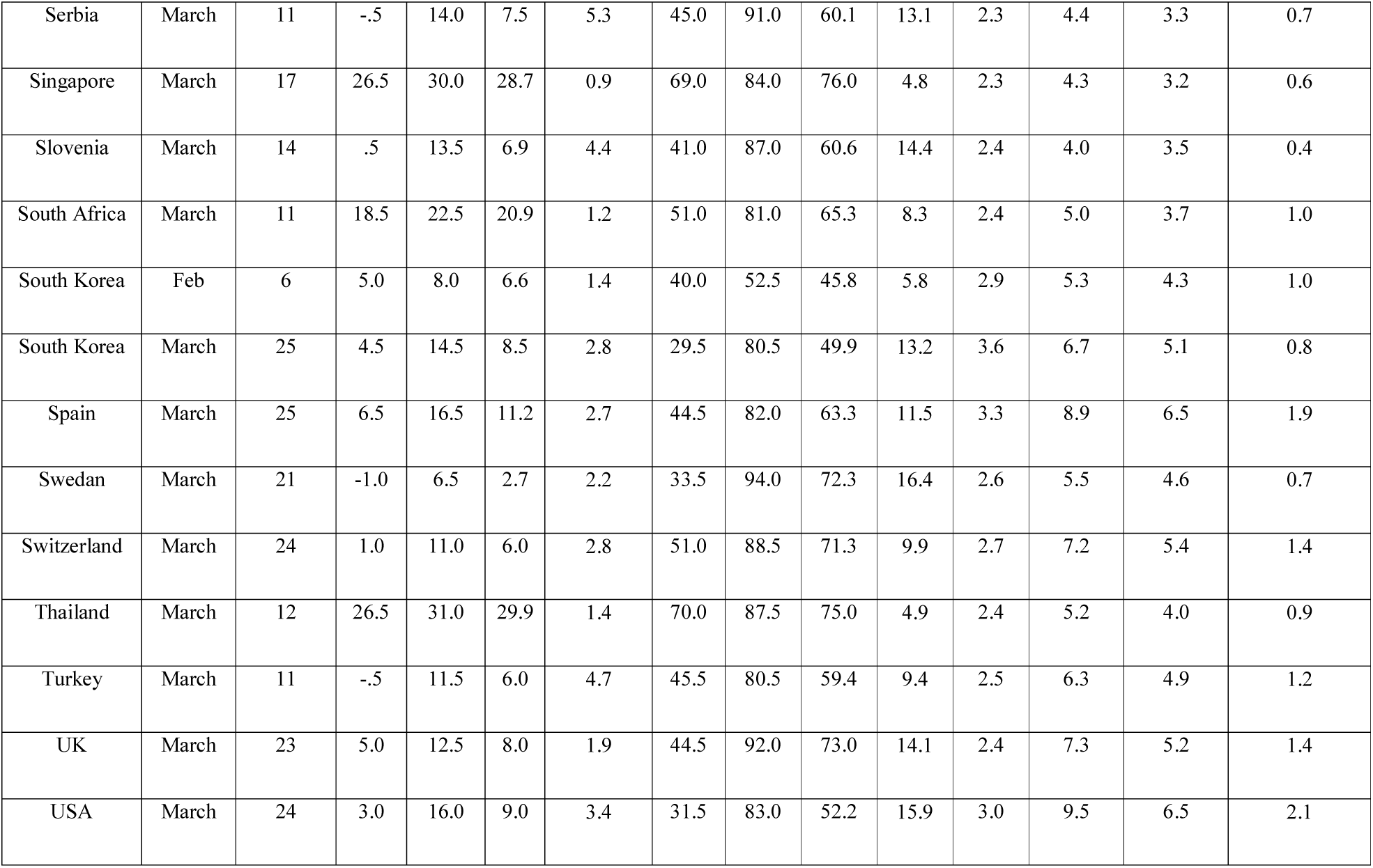
Average temperature, relative humidity and COVID-19 cases in 57 countries

